# Growth trajectories and risk of underweight and overweight in childhood cancer patients and survivors: a longitudinal multicentre cohort study protocol

**DOI:** 10.64898/2026.10.01.26364469

**Authors:** Alice Sofia Calvello, Luigi Annicchiarico, Yara Shoman, Lorenz Manuel Leuenberger, Maria José Bento, Julia Bielicki, Claudia Boettcher, Pierluigi Brazzola, Paula Breia, Gabriela Caldas, Beatrice Drexler, Daniel Drozdov, Manuel Diezi, Janez Jazbec, Markus Joerger, Daniel Konrad, Claudia E. Kuehni, António Lourenço, Christoph Saner, Katrin Scheinemann, Freimut Schilling, Milica Stefanović, Tiago Taveira Gomes, André O. von Bueren, Nicolas Waespe, Andreas Wicki, Christina Schindera, Fabiën N. Belle, the Growth consortium

**Affiliations:** Paediatric Routine Data, Childhood Cancer Research Group, Institute of Social and Preventive Medicine, University of Bern, Bern, Switzerland; Graduate School for Health Sciences, University of Bern, Bern, Switzerland; Department of Epidemiology, North Region Cancer Registry of Portugal (RORENO), Portuguese Oncology Institute of Porto (IPO Porto), Porto, Portugal; Group of Epidemiology, Outcomes, Economics and Management in Oncology – Research Centre, Porto Comprehensive Cancer Centre (Porto.CCC) & RISE@CI-IPOP (Health Research Network), Portugal; Department of Paediatric Infectious Diseases, University Children’s Hospital Basel, Basel, Switzerland; Centre for Neonatal and Paediatric Infection, St George’s University, London, United Kingdom; Division of Paediatric Endocrinology, Diabetology and Metabolism, Department of Paediatrics, Inselspital, Bern University Hospital, University of Bern, Bern, Switzerland; Paediatric Institute of Italian Switzerland, Bellinzona, Bellinzona, Switzerland; Research Center, Portuguese Oncology Institute Francisco Gentil (IPO Lisboa), Lisbon, Portugal; Paediatric Oncology Department (IPO Lisboa) and South Portugal Cancer Registry (ROR-Sul), Portuguese Oncology Institute Francisco Gentil (IPO Lisboa), Lisbon, Portugal; Department of Haematology, University Hospital Basel, Basel, Switzerland; Division of Paediatric Haematology Oncology, Children’s Hospital, Kantonsspital Aarau (KSA), Aarau, Switzerland; Paediatric Haemato-Oncology Unit, Service of Paediatrics, Lausanne University Hospital, University of Lausanne, Switzerland; Department of Paediatric Haematology and Oncology, University Children’s Hospital, University Medical Centre Ljubljana, Slovenia; Department of Medical Oncology and Haematology, Health Ostschweiz (HOCH), St. Gallen, Switzerland; Division of Paediatric Endocrinology and Diabetology, University Children’s Hospital, University of Zurich, Zurich, Switzerland; Children’s Research Centre, University Children’s Hospital, University of Zurich, Zurich, Switzerland; Zurich Centre for Integrative Human Physiology, University of Zurich, Zurich, Switzerland; Division of Paediatric Haematology/Oncology, Department of Paediatrics, Inselspital, Bern University Hospital, University of Bern, Bern, Switzerland; Epidemiology Research Unit and South Portugal Cancer Registry (ROR-Sul), Portuguese Oncology Institute Francisco Gentil (IPO Lisboa), Lisbon, Portugal; Division of Paediatric Endocrinology, Diabetology and Metabolism, Department of Paediatrics, Children’s University Hospital Bern, Julie-von-Jenner-Haus, University of Bern, Bern, Switzerland; Department of Biomedical Research, University of Bern, Bern, Switzerland; Murdoch Children’s Research Institute, The Royal Children’s Hospital, Parkville, Victoria, Australia; Division of Haematology/Oncology, Children’s Hospital of Eastern Switzerland, St. Gallen, Switzerland; Faculty of Health Sciences and Medicine, University of Lucerne, Lucerne, Switzerland; Children’s Hospital of Central Switzerland, Lucerne, Switzerland; Health Research Network, University of Porto, Porto, Portugal MTG Research and Development Lab, Porto, Portugal; Division of General Paediatrics, Paediatric Haematology and Oncology Unit, Department of Paediatrics, Gynaecology and Obstetrics, University Hospitals of Geneva, Geneva, Switzerland; CANSEARCH Research Platform for Paediatric Oncology and Haematology, Faculty of Medicine, Department of Paediatrics, Gynaecology and Obstetrics, University of Geneva, Geneva; Department of Medical Oncology and Hematology, University Hospital Zurich and University of Zurich, Zurich, Switzerland; Division of Paediatric Oncology/Haematology, University Children’s Hospital Basel, University of Basel, Basel, Switzerland

**Keywords:** Paediatric, survivorship, obesity, body mass index, linear growth, late effects, cancer treatment

## Abstract

**Background:** Underweight, overweight, and linear growth during and after cancer treatment can impact treatment efficacy and survival in childhood cancer patients, and long-term health in childhood cancer survivors (CCS). However, scientific evidence on the weight status and growth of CCS is limited. Previous studies have been small, focused on specific cancer types, or lacked objectively measured longitudinal data. The availability of standardized, semantically interoperable routine data from oncology clinics offers an unprecedented opportunity to investigate nutritional status across all childhood cancer types, treatment regimens, and critical time windows.

**Objectives:** We will investigate underweight, overweight, and growth trajectories from birth to adulthood in childhood cancer patients and survivors. We will identify predictors associated with distinct growth and weight trajectories and assess the consistency of findings across European countries.

**Methods:** The Growth study will collect routinely recorded sociodemographic, clinical, and anthropometric data from paediatric and adolescent cancer patients (aged <20 years) from Switzerland, Portugal and Slovenia, We will include patients diagnosed between 2004-2026, by linking data from the Swiss Childhood Cancer Registry (ChCR), the Swiss Childhood Cancer Survivor Study (SCCSS), the Swiss Federal Statistical Office (FSO), and oncology clinics in Switzerland.

Deterministic linkage of data sources (ChCR, SCCSS, oncology clinics, and FSO) will be performed using social security numbers. We will calculate age- and sex-specific z-scores for weight, height, and BMI based on country-specific and World Health Organization growth references. Latent trajectory modelling will be applied to identify distinct growth and weight trajectories and to detect subgroups at increased risk of growth impairment. We will use logistic regression models to identify predictors of impaired growth.

**Results:** The project was funded in 2025; first study results are expected to be published by autumn 2027. We will obtain routinely collected data from approximately 12,250 childhood cancer patients and survivors - 7,800 from Switzerland, 3,350 from Portugal, and 1,100 from Slovenia.

**Conclusions:** The Growth study will provide insights into the nutritional status of childhood cancer patients and survivors and improve understanding of growth patterns in this population. Identifying high-risk groups and tumour- or treatment-related factors will inform targeted surveillance strategies and early nutritional interventions.

## INTRODUCTION

The World Health Organization (WHO) recommends regular monitoring of height, weight, and BMI in children with cancer to guide personalised nutritional care, including timely nutritional interventions and education^1^. Both underweight and overweight are common during and after childhood cancer treatment, reflecting the impact of cancer and its treatment on growth and development during childhood^2^. Underweight during treatment has been reported in up to 70% of patients, depending on cancer and treatment type^3^, and may persist long after treatment. This is particularly relevant for those diagnosed at a young age, with underweight at diagnosis, chronic conditions, or coming from a lower socioeconomic background^4,5^. Overweight is also common, driven by glucocorticoid exposure, endocrine changes, reduced physical activity, and excess nutritional intake, with persistent weight gain particularly affecting survivors of leukaemia, lymphoma, and with a (supra)sellar tumour^3,6–11^. Cancer and its treatment can also impair linear growth through endocrine late effects such as growth hormone deficiency, hypothyroidism, and hypogonadism, especially in patients treated with cranial or total body irradiation^12–19^. Although some survivors achieve normal adult height, others experience persistent growth deficits into adulthood^20,21^.

Many risk factors for growth and weight disorders during treatment are well established, but evidence of their impact on long-term outcomes in childhood cancer survivors—such as overweight, underweight, and short stature—remains scarce. Additionally, only a few longitudinal studies have modelled growth trajectories using repeated measurements of height and weight in this population^22,23^. Last, studies are mainly conducted in cohorts from North America, limiting the generalisability of results to different settings^4,17,24^.

Electronic health records (EHRs) offer an opportunity to address these knowledge gaps by providing routinely collected longitudinal data from clinical practice. Although primarily collected for administrative and clinical care purposes, EHRs contain detailed information on patients’ diagnoses, treatments, demographic characteristics, and repeated anthropometric measurements, making them a valuable resource for studying long-term growth and weight outcomes. Such records can be used for large-scale, multi-institutional research and longitudinal analyses^25^; they can also be linked to different data sources and registries via data linkages. The collection of EHRs from multiple institutions and their linkage with additional data guarantees a considerable sample size and a long follow-up. Multiple initiatives are currently using EHRs for clinical research in Switzerland, with the aim of providing a safe, interoperable, and shareable network, like the Swiss Personalized Health Network (SPHN) and Digisanté^26,27^. Initiatives for safe and systematic digitalisation of health-related data are also conducted in Portugal, with the establishment of the Electronic Health Registry (Registo de Saúde Eletrónico), a national platform which centralizes clinical information^28^; similarly, a project to digitalise health data was completed in Slovenia (eZdravje), to introduce effective and user-friendly digital solutions into the Slovenian healthcare system and to apply established semantic and technical standards that enable the interoperability of various local information systems^29^.

Therefore, we will make use of EHRs linked with cancer and population registries to obtain a rich dataset, with a high sample size, a long follow-up, and with multiple measurements from patients and survivors diagnosed with different cancer types. We will investigate underweight, overweight, and linear growth of childhood cancer patients and survivors from birth to adulthood across three European countries (Switzerland, Portugal and Slovenia). We will examine associated predictors of growth and assess the consistency of our findings across countries.

## METHODS

### Study design

The Growth study is a retrospective multicentre cohort study using EHR data collected during cancer treatment and follow-up care over more than 20 years to investigate longitudinal anthropometric trajectories. The study includes patients diagnosed with leukaemia, lymphoma, central nervous system tumours, or malignant solid tumours before the age of 20 years between 2004-2026 who were treated at one of the participating oncology clinics. Inclusion of Portuguese patients is limited to those diagnosed since 2012 at Instituto Português de Oncologia do Porto Francisco Gentil (IPO-PORTO), or since 2017 at Instituto Português de Oncologia de Lisboa Francisco Gentil (IPOLFG), while inclusion of Slovenian patients is limited to those diagnosed since 2011 at the Paediatric Oncology Clinic, University Medical Centre Ljubljana (UMCL). Height and weight are routinely measured during clinical care, resulting in repeated anthropometric measurements for most patients. During treatment, measurements are typically obtained weekly or biweekly^30^. After treatment, follow-up visits are generally scheduled every 1–3 months during the first year, every 3–6 months for the following 5 years, and annually thereafter, or according to the patient’s medical history^31^. During each follow-up visit health care professionals normally measure the patient’s height and weight.

The study is registered at ClinicalTrials.gov (identifier: NCT07750444).

### Data sources

The Growth study will use and link patient data from different sources (Table 1).

**Table 1.** Data available from each data providers as part of the Growth study.

| Data source | Variables list | Availability |
| --- | --- | --- |
| **Switzerland** |  |  |
| Swiss Childhood Cancer Registry (ChCR) | • Demographic: sex, date of birth, place of birth, nationality etc.<br>• Diagnosis: date of incidence, classification, laterality, stage, tumour size, diagnostic institute, etc.<br>• Second primary neoplasms<br>• Patient's health: inherited predispositions, prior diseases, comorbidities, late effects<br>• Anthropometry: height, weight<br>• Treatment: study participation (protocol, regimen), chemotherapy, radiotherapy, surgery, hematopoietic stem cell transplantation, start/end date<br>• Remission status<br>• Socioeconomic: Swiss socioeconomic position (Swiss-SEP) at diagnosis | All patients diagnosed since 1976 |
| Swiss oncology clinics (Aarau, Bern, Basel, Geneva, Lausanne, Luzern, St. Gallen, and Zurich) | • Anthropometry: repeated heights and weights, dates<br>• Medication: chemotherapy components, targeted cancer therapies including immunotherapies, dosage, date of prescription/administration<br>• Surgeries: Swiss classification of surgical interventions (CHOP) | Available anthropometric data since 2004 |
| Swiss Federal Statistical Office | • Child: gestational age, nationality, vital status, date<br>• Birth: single or multiple birth, type of birth, birth order<br>• Anthropometric data: birth weight, birth length<br>• Parents: age, nationality, profession, civil status (mother)<br>• Socioeconomic: Swiss socioeconomic position (Swiss-SEP) at birth | National data available since 1969 |
| Swiss Childhood Cancer Survivor Study (SCCSS) | • Anthropometric data: self-reported height, weight<br>• Health behaviour (diet, physical activity, smoking, alcohol), quality of life, puberty, health (growth hormone deficiencies, hyper-/hypopituitarism, diabetes) | Patients diagnosed since 1976 |
| **Portugal** |  |  |
| Portuguese Institute of Oncology of Porto and Lisbon (IPO-Porto, IPOLFG) | • Demographic: sex, date of birth, place of birth, nationality etc.<br>• Diagnostic: date of diagnosis, tumour type, tumour stage<br>• Anthropometry: repeated heights and weights, dates of measurement<br>• Treatment: study participation (protocol, regimen), chemotherapy, radiotherapy, surgery, hematopoietic stem cell transplantation, start/end date<br>• Remission status | Anthropometric data available since 2012 (IPO-Porto) or 2017 (IPOLFG) |
| **Slovenia** |  |  |
| University Medical Centre Ljubljana (UMCL) | • Demographic: sex, date of birth, place of birth, nationality etc.<br>• Diagnostic: date of diagnosis, tumour type, tumour stage<br>• Anthropometry: repeated heights and weights, dates of measurement<br>• Treatment: study participation (protocol, regimen), chemotherapy, radiotherapy, surgery, hematopoietic stem cell transplantation, start/end date<br>• Remission status<br>• Socioeconomic status indicator | Anthropometric data available since 2011 |

Abbreviations: Swiss Childhood Cancer Registry (ChCR), electronic health records from paediatric oncology clinics, the Swiss Federal Statistical Office (FSO), Swiss Socioeconomic position index (Swiss-SEP<sup>33</sup>), the Swiss Childhood Cancer Survivor Study (SCCSS), the Portuguese Institute of Oncology of Porto and Lisbon (IPO-Porto, IPOLFG), the University Medical Centre Ljubljana (UMCL).

#### Switzerland

##### The Childhood Cancer Registry in Switzerland (ChCR)

The ChCR is a national, population-based cancer registry established in 1976^32^. It registers all patients diagnosed with cancer under the age of 20 who are residents in Switzerland at the time of diagnosis, and provides detailed clinical information, including tumour characteristics and treatments.

##### Swiss oncology clinics

The Swiss oncology clinics will provide routinely collected anthropometric information from EHRs of all inpatient and outpatient visits of childhood cancer patients since 2004. The nine paediatric oncology clinics are located in Aarau, Basel, Bellinzona, Bern, Geneva, Lausanne, Lucerne, St. Gallen, and Zurich; the three participating adult oncology clinics are in Basel, St. Gallen, and Zurich. We will request data on chemotherapy, their dosages, date of prescription/administration, and height and weight measurements for dosage calculations from all clinics that use Computer Aided Therapy for Oncology (BD CATO™) software, an integrated software system for managing oncology medication workflows or similar. Other medication information, such as growth hormone treatment and thyroid medication, will be available. Oncology clinics will provide Swiss Classification of Operations (CHOP) procedure codes originally based on International Classification of Diseases (ICD-9-CM) codes, diagnosed late effects coded by International Classification of Diseases, 10th revision (ICD-10), and nutritional interventions including oral nutritional supplements, enteral and parenteral nutrition making use of the Anatomical Therapeutic Chemical (ATC) classification.

##### The Swiss Federal Statistical Office (FSO)

The FSO has recorded all live births and deaths in Switzerland since 1969 in its Vital Statistics (BEVNAT) registry (www.bfs.admin.ch). Data collected after 2010 can be linked using the patient’s social security numbers known as the Old-Age and Survivors’ Insurance (OASI) numbers, and can provide birth weight, length, gestational age, nationality, mortality data, and the Swiss neighbourhood index of socioeconomic position (Swiss-SEP), an area-based socioeconomic index based on neighbourhood summary data on education, occupation, rent, and overcrowding from national censuses, which is derived from a patient’s home address^33^.

##### The Swiss Childhood Cancer Survivor Study (SCCSS)

The SCCSS invites all children and adolescents registered in the ChCR residing in Switzerland at the time of diagnosis and having survived for at least five years^34^. Data include self-reported heights and weights from more than 4,050 participants diagnosed with childhood cancer between 1976 and 2019. The SCCSS instructs all study participants (baseline and follow-up questionnaires) to record height without shoes and weight without clothes. Furthermore, the SCCSS collects detailed participant information on health behaviours (including physical activity, screen time, smoking habits, diet, and alcohol consumption), clinical conditions (including cardiovascular and endocrine problems, musculoskeletal impairments), living situation, and quality of life.

#### Portugal

##### Oncology clinics in Portugal

In Portugal, two clinics will be involved: Instituto Português de Oncologia do Porto Francisco Gentil – IPO-PORTO and Instituto Português de Oncologia de Lisboa Francisco Gentil – IPOLFG. The diagnoses identified in these clinics account for approximately 65% of all the childhood cancer cases in the country.

Diagnosis, anthropometric and treatment data will be available for analysis for children and adolescents diagnosed with cancer under the age of 19, who received care at one of the institutes since 2012.

#### Slovenia

##### Oncology clinics in Slovenia

Slovenia has one single paediatric oncology clinic, namely the Slovenia University Medical Centre Ljubljana (UMCL). Diagnosis, anthropometric, and treatment data will be available for patients diagnosed before 19 years, treated since 2011.

### Data Linkage

We will link EHRs from Swiss oncology clinics with the ChCR, the SCCSS questionnaire study^34^, and with routine statistics of the FSO (https://www.bfs.admin.ch/bfs/en/home.html). Data will be linked using deterministic linkage using OASI numbers.

### Data platform and Governance structure

Data from Switzerland and Slovenia will be accessible on BioMedIT (https://www.biomedit.ch/), a national secure computing network for collecting, storing, and analysing health-related data^35^. Access to the workspace will be coordinated by permission managers and restricted to permitted users. Users must perform the mandatory data protection and IT security awareness training and accept the terms of use. Secure and encrypted modalities will be used for data transfers between oncology clinics and BioMedIT, and between external data providers and BioMedIT. Portuguese data will be analysed following an “algorithm-goes-to-data” framework performed by MTG Research and Development Lab, a private research and consulting company specialising in real-world health data (https://mtg.pt/). The analysis code will be sent for execution and run locally on the oncology clinics’ servers of IPO-Porto and IPO-LFG. Aggregated results will be extracted and sent to BioMedIT.

The data transfers are captured in a consortium agreement including a project description, governance, data transfer and use agreement and data transfer and processing agreement, security policy, and authorship guidelines. The governance structure of the Growth study consists of an Executive Board including a representative of each party. The Executive Board is the highest decision-making body deciding on requests for reuse of data, admission of new parties, and the fate of the data at the end of the project.

### Ethics

The Ethics Committee of the Canton of Bern, Switzerland (KEK Bern), approved the Growth study in 2026 (2026-00308). The Commission for Professional Ethical Issues at UMCL approved the Growth Study in Slovenia (KSEV-4-070726), while a country-specific ethical approval request is ongoing in Portugal. Ethical approval of the SCCSS survey was granted by the Ethics Committee of the Canton of Bern (KEK-BE: 166/2014 and 2021-01462).

### Statistical Analyses

All eligible patients will contribute person-time from cohort entry (diagnosis or start of follow-up, as applicable) until death, loss to follow-up, withdrawal, or end of study. Thus, patients who die during follow-up contribute data up to their date of death according to their accrued person-time.

We will calculate age- and sex-dependent z-scores for height, weight, and BMI based on country-specific growth references and the World Health Organization (WHO) growth references using Cole’s Lambda-Mu-Sigma (LMS) method^36–38^. We will detect outliers resulting from recording or transcription errors using the method from Daymont et al. and will re-centre height, weight, and BMI z-scores around the median at each age to account for the possibility that the study sample differs from the reference population^39^. We will exclude biologically implausible values in longitudinal height and weight measurements using the *growthcleanr* algorithm^39^. We will use cutoff values for BMI published by the WHO and country-specific growth references to classify children^40^. For adults, we will use the WHO cutoff values^41^. We will fit linear, logistic, and multinomial mixed-effects models to evaluate factors associated with overweight, underweight, and linear growth at different time points. We will perform separate analyses for the most frequent childhood cancer types, e.g., acute lymphoid leukaemia and central nervous system tumours.

We will apply generalised additive models (GAMs) and group-based trajectory modelling (GBTM) to estimate growth and BMI trajectories, to identify subgroups, and to determine critical periods of weight loss, weight gain, or impaired growth within each class and overall. We will use Bayesian Information Criterion (BIC) to choose the number of classes, while also ensuring clear class separation (entropy near 1.0), sufficient sample size per trajectory (>5%), and clinical relevance^42^. We anticipate identifying 4–6 BMI trajectories and 3–4 linear growth trajectories^43,44^. To identify predictors of trajectory membership, we will use trajectory membership as a categorical value in country-specific, multinomial logistic regression models, which will account for sociodemographic, early life, and clinical factors, such as cancer type and treatments. For trajectory modelling, individuals with fewer than three measurements of height and weight will be excluded from the analysis.

We will conduct parallel analyses in the three independent cohorts (Switzerland, Portugal, and Slovenia) to assess the consistency of the findings across countries.

A description of missing values for disease and treatment data will be conducted, with the aim of observing patterns of missingness and evaluating whether missingness can be considered at random or not. When appropriate, multiple imputation will be performed for handling missing data.

## RESULTS

We will collect and analyse clinical, anthropometric, demographic and socioeconomic data of approximately 12,250 patients. Of these, approximately 7,800 will be from Switzerland, 3,350 from Portugal, and 1,100 from Slovenia (Table 2). For the Swiss cohort, the linkage with the SCCSS questionnaire study will additionally provide repeated self-reported heights and weights of more than 4,050 survivors who received the questionnaire at least 5 years after diagnosis. We estimate that most patients have at least 30 anthropometric assessments available at birth (n=1), during treatment (n>16), during follow-up (n>13).

**Table 2.** Expected number of childhood cancer patients by type of tumour and country (Switzerland: 2004 – 2026*; Slovenia: 2011 – 2026*; Portugal: 2012 – 2026*).

| ICCC-3<br>main group | Diagnosis | Switzerland | Portugal | Slovenia |
| --- | --- | --- | --- | --- |
| I | Leukaemias | 1769 | 775 | 323 |
| II | Lymphomas | 796 | 637 | 157 |
| III | CNS neoplasms | 1448 | 617 | 254 |
| IV | Neuroblastoma | 400 | 85 | 54 |
| V | Retinoblastoma | 700 | 40 | 16 |
| VI | Renal tumours | 242 | 101 | 30 |
| VII | Hepatic tumours | 84 | 56 | 4 |
| VIII | Malignant bone tumours | 308 | 199 | 81 |
| IX | Soft tissue sarcomas | 466 | 195 | 30 |
| X | Germ cell tumours | 202 | 105 | 52 |
| XI & XII | Other tumours | 1404 | 540 | 127 |
| **TOTAL** |  | **7819** | **3355** | **1132** |

Abbreviations: Central Nervous System (CNS), International Classification of Childhood Cancer, third edition (ICCC-3) \*Diagnoses to 2025 and 2026 are based on data from previous years

We anticipate the first results of the Growth study to be available by 2027.

## DISCUSSION

This study represents one of the first European longitudinal cohorts with up to 20 years of follow-up across three countries, focusing on underweight, overweight, and growth of childhood cancer patients and survivors. Optimal nutritional status is a key component of paediatric oncology care, as emphasised by the WHO^45^. Malnutrition—encompassing both undernutrition and overnutrition—is common during and after treatment and is associated with increased risks of infection, treatment-related toxicity, and reduced survival^46,47^. As a potentially modifiable risk factor, nutritional status represents a possible target for early intervention to improve long-term health outcomes and quality of life^46^. Childhood is a critical period for the development of excess weight, which often tracks into adulthood^48,49^. However, it remains unclear if growth trajectories of childhood cancer patients and survivors are comparable with the growth references for the general paediatric population.

Evaluating growth patterns and risk factors across different clinics and countries is challenging due to variations in the collection of EHRs, as well as differences in population characteristics and lifestyle contexts. Moreover, planning randomized clinical trials in this population of patients is often impractical and unethical: in general, clinical trials - the gold standard to establish causal relationships between exposures and outcomes - are expensive and burdensome for both patients and treatment centres; this is particularly true for childhood cancer patients, who already participate in trials related to treatment efficacy. Therefore, this study aims to compare longitudinal growth and nutritional trajectories across multiple European settings to disentangle the relative contributions of country-specific versus tumour- and treatment-related factors. It will help to identify areas of need and to prioritise future trials.

### Strengths and limitations

This study has several strengths: the use of data from Switzerland, Portugal, and Slovenia will allow for a large sample size and a broad diagnostic coverage. The linkages between many different Swiss sources will provide a rich and high-quality dataset, allowing for a thorough examination of exposures throughout the different phases of growth and development. This study will primarily utilise objective measurements of height and weight recorded by health professionals from birth through follow-up care. Additionally, the long follow-up period will allow for the investigation of late effects on height and weight. Conversely, the inclusion of clinics from multiple countries, each with differences in clinical and data collection practices, introduces heterogeneity that may reduce the interpretability of our results. In addition, because electronic health records are collected primarily for administrative purposes, they contain a limited number of extractable variables and often include unstructured information. Missing or inaccurately coded data, variability in documentation practices, incomplete capture of care received outside the healthcare system, and the inability to account for all potential confounders may introduce bias.

### Outlook

This study will harness standardised, accessible, and interoperable health data to advance research on childhood cancer, underweight, overweight, and linear growth in Switzerland and establish key international collaborations.

This unique dataset, covering anthropometric data from birth to late survivorship, will provide crucial insights into nutritional status in childhood cancer patients and survivors, as growth management is essential for optimising physical, cognitive, and emotional development and ensuring long-term health. By providing a comprehensive overview of BMI and growth patterns and their predictors across different cancer types—all rare diseases—this study will identify tumours, treatments, and critical time windows of concern. Vulnerable individuals requiring close follow-up care could be identified and trajectories associated with suboptimal growth outcomes could be highlighted. Our findings will inform clinical practice and improve patient care in a research area where randomised trials are often impractical.

**Figure 1.**
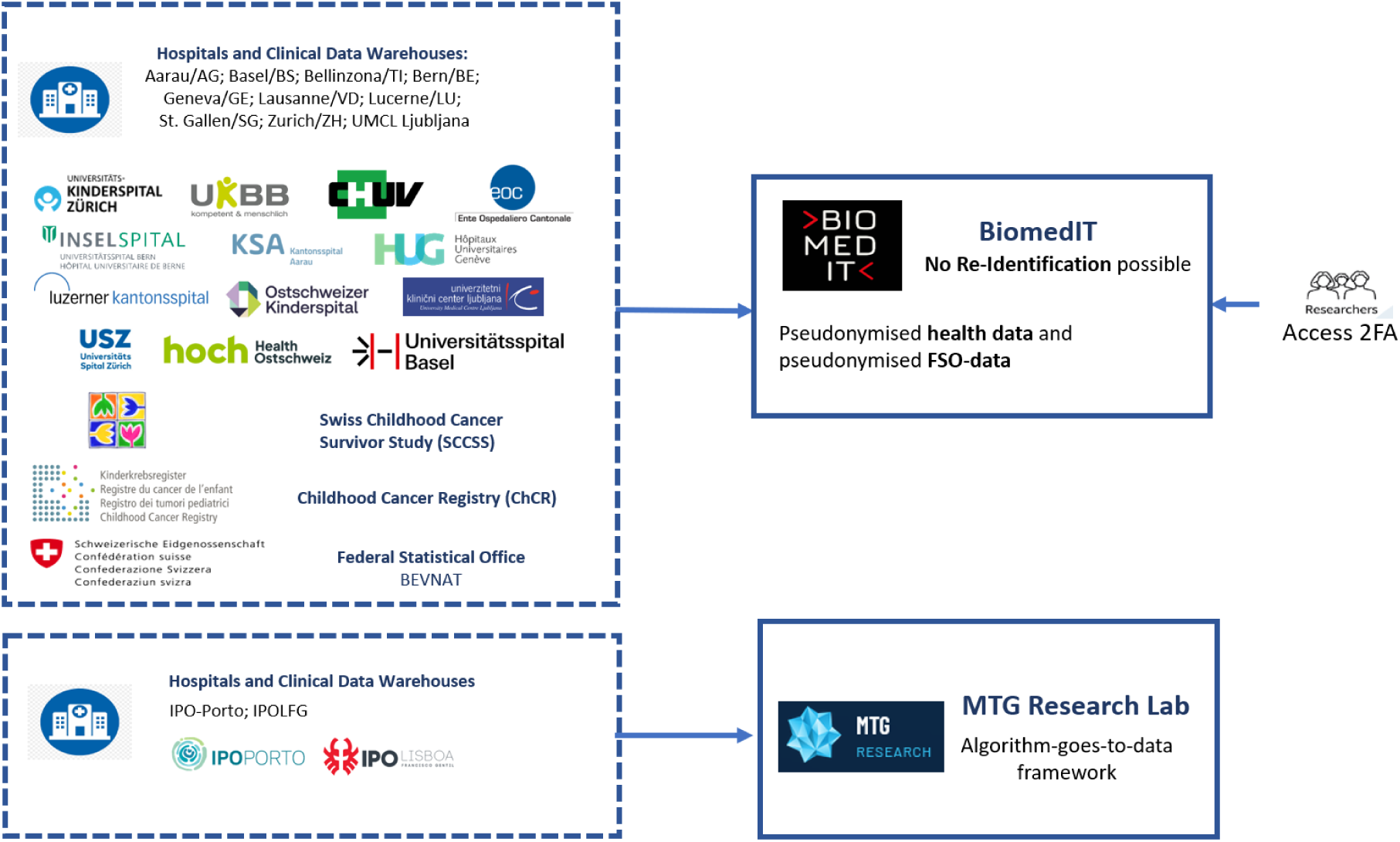
Overview of data sources and data management processes within the Growth study. Abbreviations: BEVNAT, Swiss Vital Statistics Registry; ChCR, Swiss Childhood Cancer Registry; FSO, Swiss Federal Statistical Office; IPOLFG, Portuguese Institute of Oncology of Lisbon; IPO-Porto, Portuguese Institute of Oncology of Porto; SCCSS, Swiss Childhood Cancer Survivors Study; UMCL, University Medical Centre Ljubljana.

## ACKNOWLEDGEMENTS

The Growth consortium includes the following:

- Luigi Annicchiarico, Institute of Social and Preventive Medicine (ISPM), University of Bern, Bern, Switzerland;
- Daniela Antunes, Health Research Network, University of Porto, Porto, Portugal MTG Research and Development Lab, Porto, Portugal;
- Fabiën Belle, Institute of Social and Preventive Medicine (ISPM), University of Bern, Bern, Switzerland;
- Julia Bielicki, Paediatric Research Center, University l Children’s Hospital Basel (UKBB), University of Basel, Basel, Switzerland & Centre for Neonatal and Paediatric Infection, St George’s, University of London, London, UK;
- Pierluigi Brazzola, Paediatric Institute of Italian Switzerland, Bellinzona, Bellinzona, Switzerland;
- Claudia Brito, Epidemiology Research Unit and South Portugal Cancer Registry (ROR-Sul), Portuguese Oncology Institute Francisco Gentil (IPO Lisboa), Lisbon, Portugal;
- Maria José Bento, Department of Epidemiology, North Region Cancer Registry of Portugal (RORENO), Portuguese Oncology Institute of Porto (IPO Porto), Porto, Portugal;
- Paula Breia, Research Center, Portuguese Oncology Institute Francisco Gentil (IPO Lisboa), Lisbon, Portugal;
- André von Bueren, Division of General Paediatrics, Paediatric Haematology and Oncology Unit, Department of Paediatrics, Gynaecology and Obstetrics, University Hospitals of Geneva, Geneva, Switzerland &CANSEARCH Research Platform for Paediatric Oncology and Haematology, Faculty of Medicine, Department of Paediatrics, Gynaecology and Obstetrics, University of Geneva, Geneva, Switzerland;
- Gabriela Caldas, Paediatric Oncology Department (IPO Lisboa) and South Portugal Cancer Registry (ROR-Sul), Portuguese Oncology Institute Francisco Gentil (IPO Lisboa), Lisbon, Portugal;
- Alice Sofia Calvello, Institute of Social and Preventive Medicine (ISPM), University of Bern, Bern, Switzerland;
- Filipa Caseiro, Paediatric Oncology Department (IPO Lisboa), Portuguese Oncology Institute Francisco Gentil (IPO Lisboa), Lisbon, Portugal;
- Helena Correia, Epidemiology Research Unit and South Portugal Cancer Registry (ROR-Sul), Portuguese Oncology Institute Francisco Gentil (IPO Lisboa), Lisbon, Portugal;
- Manuel Diezi, Paediatric Haemato-Oncology Unit, Service of Paediatrics, Lausanne University Hospital, University of Lausanne, Switzerland;
- Beatrice Drexler, Department of Hematology, University Hospital Basel, Basel, Switzerland;
- Daniel Drozdov, Division of Paediatric Haematology Oncology, Children’s Hospital, Kantonsspital Aarau (KSA), Aarau, Switzerland;
- Teresa Garcia, Department of Epidemiology, North Region Cancer Registry of Portugal (RORENO), Portuguese Oncology Institute of Porto (IPO Porto), Porto, Portugal;
- Janez Jazbec, Department of Paediatric Haematology and Oncology, University Children’s Hospital, University Medical Centre Ljubljana, Slovenia;
- Markus Joerger, Department of Medical Oncology and Hematology, Health Ostschweiz (HOCH), St. Gallen, Switzerland;
- Daniel Konrad, Division of Paediatric Endocrinology and Diabetology, University Children’s Hospital, University of Zurich, Zurich, Switzerland & Children’s Research Centre, University Children’s Hospital, University of Zurich, Zurich, Switzerland & Zurich Centre for Integrative Human Physiology, University of Zurich, Zurich, Switzerland;
- Primož Kotnik, Department of Paediatric Haematology and Oncology, University Children’s Hospital, University Medical Centre Ljubljana, Slovenia;
- Claudia Kuehni, Institute of Social and Preventive Medicine (ISPM), University of Bern, Bern, Switzerland & Division of Paediatric Haematology/Oncology, Department of Paediatrics, Inselspital, Bern University Hospital, University of Bern, Bern, Switzerland;
- António Lourenço, Epidemiology Research Unit and South Portugal Cancer Registry (ROR-Sul), Portuguese Oncology Institute Francisco Gentil (IPO Lisboa), Lisbon, Portugal;
- Rafael Luis, Research Center, Portuguese Oncology Institute Francisco Gentil (IPO Lisboa), Lisbon, Portugal;
- Rita Luz, Health Research Network, University of Porto, Porto, Portugal MTG Research and Development Lab, Porto, Portugal;
- Alexandra Mayer, Epidemiology Research Unit and South Portugal Cancer Registry (ROR-Sul), Portuguese Oncology Institute Francisco Gentil (IPO Lisboa), Lisbon, Portugal;
- Carla Pereira, Research Center, Portuguese Oncology Institute Francisco Gentil (IPO Lisboa), Lisbon, Portugal;
- Ana Sà Sousa, Health Research Network, University of Porto, Porto, Portugal MTG Research and Development Lab, Porto, Portugal;
- Neza Salobir, Department of Paediatric Haematology and Oncology, University Children’s Hospital, University Medical Centre Ljubljana, Slovenia;
- Christoph Saner, Division of Paediatric Endocrinology, Diabetology and Metabolism, Department of Paediatrics, Children’s University Hospital Bern, Julie-von-Jenner-Haus, University of Bern, Bern, Switzerland & Department of Biomedical Research, University of Bern, Bern, Switzerland & Murdoch Children’s Research Institute, The Royal Children’s Hospital, Parkville, Victoria, Australia;
- Katrin Scheinemann, Division of Haematology/Oncology, Children’s Hospital of Eastern Switzerland, St. Gallen, Switzerland & Faculty of Health Sciences and Medicine, University of Lucerne, Lucerne, Switzerland;
- Freimut Schilling, Children’s Hospital of Central Switzerland, Lucerne, Switzerland;
- Christina Schindera, Institute of Social and Preventive Medicine (ISPM), University of Bern, Bern, Switzerland & Division of Paediatric Oncology/Haematology, University Children’s Hospital Basel, University of Basel, Basel, Switzerland;
- Yara Shoman, Institute of Social and Preventive Medicine (ISPM), University of Bern, Bern, Switzerland;
- Grit Sommer, Institute of Social and Preventive Medicine (ISPM), University of Bern, Bern, Switzerland;
- Ben Spycher, Institute of Social and Preventive Medicine (ISPM), University of Bern, Bern, Switzerland;
- Milica Stefanović, Department of Paediatric Haematology and Oncology, University Children’s Hospital, University Medical Centre Ljubljana, Slovenia;
- Tiago Taveira-Gomes, Health Research Network, University of Porto, Porto, Portugal MTG Research and Development Lab, Porto, Portugal;
- Nicolas Waespe, Division of Paediatric Haematology/Oncology, Department of Paediatrics, Inselspital, Bern University Hospital, University of Bern, Bern, Switzerland & CANSEARCH Research Platform for Paediatric Oncology and Haematology, Faculty of Medicine, Department of Paediatrics, Gynaecology and Obstetrics, University of Geneva, Geneva;
- Andreas Wicki, Department of Medical Oncology and Hematology, University Hospital Zurich and University of Zurich, Zurich, Switzerland.

## AUTHORS’ CONTRIBUTIONS

Conceptualization: FNB, CS

Funding acquisition: FNB, CS

Methodology: ASC, LA, YS, FNB, CS, CEK

Project administration: FNB

Supervision: FNB

Validation: all authors

Writing – original draft: ASC, LA, YS, FNB

Writing – review & editing: all authors

## Data availability

The Growth study has a policy of collaborative research and is open for data sharing. Data can be made available for researchers fulfilling all ethical and legal requirements; detailed information is available on the SCCSS website: https://www.swiss-ccss.ch/en/research-projects/. Requests for access to the data or collaboration can be addressed to the principal investigators.

